# Changes in antidepressant use in Australia: A nationwide analysis prior to and during the COVID-19 pandemic (2015-2021)

**DOI:** 10.1101/2021.11.24.21266837

**Authors:** Juliana de Oliveira Costa, Malcolm B. Gillies, Andrea L. Schaffer, David Peiris, Helga Zoega, Sallie-Anne Pearson

## Abstract

**Background:** Depression and anxiety affect 4% to 14% of Australians every year; symptoms may have been exacerbated during the COVID-19 pandemic. We examined recent patterns of antidepressant use in Australia in the period 2015 to 2021, which includes the first year of the pandemic.

**Methods:** We used national dispensing claims for people aged ≥10 years to investigate annual trends in prevalent and new antidepressant use (no antidepressants dispensed in the year prior). We conducted stratified analyses by sex, age group and antidepressant class. We report outcomes from 2015 to 2019 and used time series analysis to quantify changes during the first year of the COVID-19 pandemic (March 2020 to February 2021).

**Results:** In 2019 the annual prevalence of antidepressant use was 170.4 per 1,000 women and 101.8 per 1,000 men, an increase of 7.0% and 9.2% from 2015, respectively. New antidepressant use also increased for both sexes (3.0% for women and 4.9% for men) and across most age groups, particularly among adolescents (aged 10-17 years; 46%-57%). During the first year of the COVID-19 pandemic, we observed higher than expected prevalent use (+2.2%, 95%CI 0.3%, 4.2%) among females, corresponding to a predicted excess of 45,217 (95%CI 5,819, 84,614) females dispensed antidepressants. The largest increases during the first year of the pandemic occurred among female adolescents for both prevalent (+11.7%, 95%CI 4.1%, 20.5%) and new antidepressant use (+15.6%, 95%CI 8.5%, 23.7%).

**Conclusion:** Antidepressant use continues to increase in Australia overall and especially among young people. We found a differential impact of the COVID-19 pandemic in treated depression and anxiety, greater among females than males, and greater among young females than other age groups, suggesting an increased mental health burden in populations already on a trajectory of increased use of antidepressants prior to the pandemic. Reasons for these differences require further investigation.

## Introduction

Australia ranks highly among OECD countries in terms of antidepressant use ^1^, with approximately 7% of adults estimated to be using antidepressants each day ^2^. Prevalence of antidepressant use has been rising for decades, in parallel with increased diagnosis of mental disorders, improved access to treatment, better tolerability of pharmacotherapies, expanding indications for use, and longer treatment durations ^3-5^.

Antidepressants are the most common medicines for treating depression and anxiety, conditions that affect about 4% and 14% of Australians each year, respectively ^6^. The prevalence of diagnosed anxiety and depression are 1.6-1.7 times more common among women than men across all age groups ^6^. Consequently, women use antidepressants at a rate 2-fold higher than men, with the highest use among older women aged 65 years or over ^7 8^. Moreover, antidepressant use increased at a higher rate among children and adolescents compared to other age groups in between 2009 and 2012 ^9 10^; this is despite concerns of increased risk of suicidal ideation and self-harm in this population ^11^. However, there are limited contemporary population-wide data on recent trends in antidepressant use, particularly among specific sub-populations.

Across the globe, the COVID-19 pandemic brought about increased levels of distress, as people experienced the fear of acquiring the infection, social isolation, food and job insecurity ^12^. Relative to other jurisdictions worldwide, Australia had low rates of COVID-19 cases in the first year of the pandemic (88.2/100,000 population), with 909 deaths as of February 28, 2021 ^13^ Nonetheless, several surveys reported that in the early months of the pandemic large proportions of adults experienced moderate to severe symptoms of depression (27%-46%) and moderate to severe symptoms of anxiety (21%-41%). These surveys also reported higher rates of depression and anxiety symptoms among women ^14-16^ and younger people ^17 18^.

Therefore, we aimed to describe recent patterns of antidepressant use in Australia, both prior to and during the first year of the COVID-19 pandemic covering the period 2015-2021. Specifically, we estimated the annual number of dispensed antidepressants, prevalent and new antidepressant use, stratified by age, sex and class. We hypothesised increased antidepressant use and that increases during the first year of the COVID-19 pandemic would be greater than predicted values.

## Methods

### Setting and access to antidepressants

Australia has a publicly funded, universal healthcare system providing subsidised medicines for all citizens and eligible residents through the Pharmaceutical Benefits Scheme (PBS). Prescription medicines for mental health treatment are primarily written by general practitioners and psychiatrists ^19^ and can be obtained in community or private hospital pharmacies ^20^.

### Medicines of interest

We included all dispensing for antidepressants, identified by the WHO Anatomical Therapeutical Classification (ATC) group N06A ^21^. We further classified the antidepressant class as selective serotonin reuptake inhibitors – SSRI, serotonin-noradrenaline reuptake inhibitors -SNRI, tricyclic antidepressants – TCA, monoamine oxidase inhibitors – MAOI, or Other antidepressants. We presented mirtazapine separately from MAOI/Other since this atypical antidepressant is commonly used as first-line treatment (See detail of codes in the appendix).

### Data sources

We used two national PBS dispensing claims data sets in this study. First, we used monthly aggregate data that counted the number of antidepressant prescriptions that were dispensed over time. These data are publicly available and are based on the actual month of dispensing; however, they provide aggregate counts only and not person-level characteristics such as age and sex. Therefore, we also used data from a 10% random sample of PBS-eligible people to estimate all individual-level patterns of antidepressant use over time. The data contain information on person-level characteristics (year of birth, sex, beneficiary status) and the dispensing of subsidised medicines (pharmacy state of dispensing, medicine dispensed, date of supply, quantity dispensed). Supply dates are offset by ±14 days to protect privacy, with the same direction of the offset for each person. The PBS data collection does not capture medicines dispensed to public hospitals inpatients or medicines privately purchased ^20^.

### Study population and characteristics evaluated

We used the PBS 10% sample data for the period between 1 January 2014 and 28 February 2021, to identify people ≥ 10 years of age dispensed antidepressants. We did not include children under 10 years due to limited use of antidepressants in this age group. At each dispensing, we categorised people by sex (male, female), prevalent versus new use, and age group: adolescents (10-17 years old), young adults (18-24 years old), adults (25-64 years old), older adults (≥ 65 years old). We defined prevalent use as a dispensing of at least one antidepressant in each calendar year and new use as a dispensing of an antidepressant with no dispensing of any antidepressants in the prior 12 months (i.e., not necessarily within the same calendar year).

### Statistical analyses

#### Patterns of antidepressant use

We used the PBS 10% sample to estimate person-level characteristics of antidepressant use. We calculated annual prevalent and new antidepressant use per 1,000 population for each calendar year using the Australian Bureau of Statistics mid-year population estimates. For new use, we adjusted the denominator by subtracting the relevant prevalent population (i.e., those who were not eligible to initiate treatment since they were already being treated). We further stratified estimates by sex and calculated results according to age and antidepressant class. Incidence estimates stratified by antidepressant class excluded individuals who had used any class of antidepressant in the preceding 12 months. We calculated the relative percentage of change over the five years before COVID-19 as: 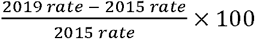

#### Changes in antidepressant use during the COVID-19 pandemic

To measure the volume of antidepressants dispensed at the national level, we used monthly aggregated PBS data, estimating the number of antidepressants as a count of items dispensed per 1,000 population per year. We then performed time series analyses using the PBS 10% sample data to evaluate the impact of the first year of the COVID-19 pandemic on prevalent and new use in Australia. We used separate analyses for prevalence and incidence and stratified by sex and age. We considered the COVID-19 period to occur from March 2020 onward since this month corresponded to the first peak of infections in Australia and the implementation of restrictions on gatherings ^13^. Our COVID-19 observation period finished in February 2021 to allow a 12-month period of observation.

We modelled trends using annual rates, to negate the need to model seasonality in dispensing and autocorrelation. This approach also dilutes the effect of the medicines stockpiling that occurred in March 2020 ^18^. To align with the 12-month COVID-19 period starting 1 March 2020, we defined each analysis year as starting on 1 March and ending on the last day (28 or 29) of February the following year. We calculated number of antidepressants dispensed, prevalence and incidence rates per 1,000 population for the analysis years 2015–2020, with adjusted denominators for incidence as described earlier. We fit linear models for the five years 2015–2019 and then predicted rates for 2020, together with 95% prediction intervals. Combined with the observed rates for 2020, this allowed us to estimate the deviations from predicted trend as percentages of the predicted rates, and to calculate excess counts by multiplying by the relevant populations. As for historical patterns of use, we stratified estimates by sex and calculated results according to age and antidepressant class.

We also performed a sensitivity analysis to account for uncertainties in the population estimates for 2020 relating to pandemic-related changes in overseas migration in and out of Australia. Estimates of the annual population growth have dropped from 1.5% for 2019 to 0.5% for 2020, mostly due to a large decrease in arrivals in the country, particularly among the younger population (15 to 29 years old). ^22^ These differences in population estimates will impact the observed and predicted rates of antidepressant use during the first year of the COVID-19 pandemic. The base case analysis used populations for 2020 with no change in population growth and the alternative case used an updated estimate that accounted for the decrease.

All analyses were performed with R version 4.0.4 (R Core Team 2021, Vienna, Austria).

### Ethics and reporting

This research was approved by the New South Wales Population and Health Services Research Ethics Committee (Approval number: 2013/11/494) and data access for the PBS 10% sample was granted by the Australian Government Services Australia External Request Committee (Approval number: MI7542).

## Results

From 2015 to 2019, prevalent antidepressant use increased for both sexes, from 159.3 to 170.4 per 1,000 females (+7.0%) and from 93.3 to 101.8 per 1,000 males (+9.2%) (Table 1). The female to male ratio was 1.7 for prevalent use across whole the study period. Prevalent antidepressant use increased with age for both sexes, being highest among people aged 85 years or older. SSRIs were the most used antidepressant class, with the prevalence among females approximately double that of males every year. SSRI prevalence increased from 83.3 to 91.8 per 1,000 females (+10.2%) and 46.5 to 53.0 per 1,000 males (+14.0%) from 2015 to 2019. Mirtazapine use also increased, while use of other/ MAOI agents declined for both sexes.

**Table 1.** Annual prevalent and new antidepressant use by demographic characteristics and medicine class, 2015 – 2019.

| Characteristics | Prevalent use per 1,000 population |  |  |  |  |  |  |  |  |  |  |  |
| --- | --- | --- | --- | --- | --- | --- | --- | --- | --- | --- | --- | --- |
|  | Females |  |  |  |  |  | Males |  |  |  |  |  |
|  | 2015 | 2016 | 2017 | 2018 | 2019 | 5-year % change | 2015 | 2016 | 2017 | 2018 | 2019 | 5-year % change |
| Overall | 159.3 | 162.5 | 164.0 | 167.3 | 170.4 | 7.0 | 93.3 | 95.4 | 96.7 | 99.3 | 101.8 | 9.2 |
| Age range |  |  |  |  |  |  |  |  |  |  |  |  |
| 10 - 17 | 39.3 | 41.4 | 43.6 | 47.4 | 50.0 | 27.2 | 24.7 | 26.1 | 28.1 | 30.8 | 33.1 | 34.0 |
| 18 - 24 | 107.6 | 113.0 | 115.1 | 118.2 | 125.7 | 16.8 | 57.2 | 60.3 | 61.9 | 62.9 | 66.4 | 16.1 |
| 25 - 34 | 116.2 | 119.8 | 120.1 | 123.1 | 127.0 | 9.3 | 71.7 | 73.4 | 74.1 | 76.4 | 78.7 | 9.8 |
| 35 - 44 | 163.4 | 164.7 | 162.0 | 162.9 | 163.3 | -0.1 | 101.1 | 103.4 | 102.8 | 103.9 | 105.1 | 4.0 |
| 45 - 54 | 193.5 | 196.2 | 199.3 | 202.8 | 205.1 | 6.0 | 114.0 | 116.3 | 117.5 | 120.8 | 123.6 | 8.4 |
| 55 - 64 | 207.4 | 209.5 | 209.9 | 213.2 | 215.0 | 3.7 | 121.8 | 123.7 | 124.9 | 127.8 | 130.6 | 7.2 |
| 65 - 74 | 221.4 | 223.6 | 226.7 | 230.5 | 233.3 | 5.4 | 126.3 | 127.5 | 129.7 | 134.4 | 137.6 | 8.9 |
| 75 - 84 | 250.6 | 253.8 | 258.3 | 262.3 | 262.7 | 4.8 | 159.3 | 160.8 | 161.1 | 163.3 | 164.9 | 3.5 |
| 85 + | 236.0 | 243.5 | 250.5 | 258.2 | 267.0 | 13.1 | 163.5 | 167.8 | 175.2 | 179.6 | 180.0 | 10.1 |
| Antidepressant class |  |  |  |  |  |  |  |  |  |  |  |  |
| SSRI | 83.3 | 85.5 | 87.5 | 89.5 | 91.8 | 10.2 | 46.5 | 48.0 | 49.6 | 51.0 | 53.0 | 14.0 |
| Mirtazapine | 11.7 | 12.2 | 12.6 | 13.3 | 13.6 | 16.2 | 10.6 | 11.3 | 11.7 | 12.2 | 12.8 | 20.8 |
| SNRI | 34.3 | 34.7 | 34.3 | 34.2 | 34.2 | -0.3 | 20.2 | 20.2 | 19.7 | 19.5 | 19.3 | -4.5 |
| TCA | 28.7 | 28.8 | 28.4 | 29.2 | 29.7 | 3.5 | 15.0 | 15.2 | 14.9 | 15.8 | 16.1 | 7.3 |
| Other/ MAOI | 1.3 | 1.2 | 1.2 | 1.1 | 1.0 | -23.1 | 0.9 | 0.8 | 0.8 | 0.8 | 0.7 | -22.2 |

Table 1. Continuation
| Characteristics | New use per 1,000 population |  |  |  |  |  |  |  |  |  |  |  |
| --- | --- | --- | --- | --- | --- | --- | --- | --- | --- | --- | --- | --- |
|  | Females |  |  |  |  |  | Males |  |  |  |  |  |
|  | 2015 | 2016 | 2017 | 2018 | 2019 | 5-year % change | 2015 | 2016 | 2017 | 2018 | 2019 | 5-year % change |
| **Overall** | 46.7 | 47.3 | 46.5 | 47.6 | 48.1 | 3.0 | 31.8 | 31.8 | 31.8 | 32.6 | 33.4 | 4.9 |
| **Age range** |  |  |  |  |  |  |  |  |  |  |  |  |
| 10 - 17 | 21.6 | 24.5 | 27.1 | 31.3 | 33.9 | 56.9 | 12.7 | 13.3 | 15.2 | 17.4 | 18.5 | 45.7 |
| 18 - 24 | 53.5 | 55.0 | 54.1 | 55.0 | 59.9 | 12.0 | 31.1 | 32.6 | 33.6 | 34.1 | 36.4 | 17.0 |
| 25 - 34 | 45.3 | 46.3 | 45.2 | 45.8 | 46.6 | 2.9 | 31.9 | 32.2 | 31.9 | 33.0 | 33.7 | 5.6 |
| 35 - 44 | 49.4 | 50.8 | 47.7 | 49.6 | 49.5 | 0.2 | 36.3 | 36.4 | 36.2 | 36.3 | 36.7 | 1.1 |
| 45 - 54 | 51.2 | 51.2 | 51.3 | 51.7 | 51.6 | 0.8 | 35.0 | 34.5 | 33.6 | 34.5 | 36.0 | 2.9 |
| 55 - 64 | 47.6 | 46.1 | 44.1 | 45.1 | 44.0 | -7.6 | 32.3 | 31.4 | 31.1 | 32.0 | 32.9 | 1.9 |
| 65 - 74 | 49.2 | 46.9 | 47.2 | 47.6 | 46.3 | -5.9 | 32.8 | 31.8 | 32.4 | 33.1 | 32.6 | -0.6 |
| 75 - 84 | 60.3 | 61.6 | 61.5 | 59.6 | 57.3 | -5.0 | 47.9 | 44.5 | 43.9 | 43.4 | 42.4 | -11.5 |
| 85 + | 60.9 | 59.4 | 58.3 | 55.9 | 54.8 | -10.0 | 52.5 | 54.8 | 52.2 | 49.0 | 46.3 | -11.8 |
| **Antidepressant class** |  |  |  |  |  |  |  |  |  |  |  |  |
| SSRI | 24.0 | 24.6 | 24.6 | 25.2 | 26.1 | 8.8 | 16.3 | 16.4 | 17.0 | 17.1 | 18.0 | 10.4 |
| Mirtazapine | 2.6 | 2.6 | 2.6 | 2.7 | 2.6 | 0.0 | 2.9 | 2.9 | 2.9 | 3.0 | 3.2 | 10.3 |
| SNRI | 3.6 | 3.3 | 2.9 | 2.8 | 2.8 | -22.2 | 2.7 | 2.4 | 2.1 | 2.0 | 1.9 | -29.6 |
| TCA | 16.5 | 16.6 | 16.3 | 16.8 | 16.6 | 0.6 | 9.8 | 9.9 | 9.7 | 10.4 | 10.2 | 4.1 |
| Other/ MAOI | 0.1 | 0.1 | 0.1 | 0.1 | 0.1 | 0.0 | 0.1 | 0.1 | 0.1 | 0.1 | 0.1 | 0.0 |

SSRI: Selective Serotonin Reuptake Inhibitors, SNRI: Serotonin-Noradrenaline Reuptake Inhibitors, TCA: Tricyclic Antidepressants, MAOI: Monoamine oxidase inhibitors

Similarly, new antidepressant use increased over the same time period for both sexes, although the increases were less pronounced than the changes in prevalence: from 46.7 to 48.1 per 1,000 females (+3.0%) and 31.8 to 33.4 per 1,000 males (+4.9%) (Table 1). Annual initiation was steady among adults aged 25 and older. In 2019, the highest overall initiation was among females aged 18 to 24 years (59.9 per 1,000), similar to estimates among older women, aged 75 to 84 years (57.3 per 1,000). However, the largest increase (+56.9%) in incident use occurred among females aged 10 to 17 years (21.6 in 2015 to 33.9 per 1,000 in 2019). Most people initiated on SSRIs, followed by TCAs. From 2015 to 2019, we observed a decrease in overall initiation of SNRIs but an increase of mirtazapine initiation among males only.

### Changes during the COVID-19 pandemic

We observed a higher than expected number of antidepressants dispensed per 1,000 population during the first year of the COVID-19 pandemic compared with the predicted based on the 2015-2019 trend (+2.8%, 95%CI 0.5%, 5.2%).

Overall, we observed marginally higher than expected prevalent antidepressant use (+1.4%, 95%CI −0.6%, 3.6%) (Table 2, Figure 1). Among females, we observed a 2.2% (95%CI 0.3%, 4.2%) increase corresponding to a predicted excess of 45,217 (95%CI 5,819, 84,614) females dispensed antidepressants during the first year of the COVID-19 pandemic. The largest increases occurred among female adolescents (+11.7%, 95%CI 2.3%, 13.2%) and young adults (+7.5%, 95%CI 2.3%, 13.2%), corresponding to an excess of 8,135 (95%CI 3,048, 13,221) and 11,910 (95%CI 3,849, 19,970) females using antidepressants during the COVID-19 pandemic, respectively. We observed changes in most antidepressant classes among females, with higher than predicted rates for SSRIs (+2.3%, 95%CI 0.7%, 4.0%) and lower than predicted rates for other/ MAOI (−6.4%, 95%CI −7.9%, −4.9%). Among males, we observed higher than expected prevalent SNRI use (+2.2%, 95%CI 0.1%, 4.2%).

**Table 2.** Changes in antidepressant use during the first year of COVID-19* according to sex, age, and antidepressant class.

| Characteristics | Prevalent use |  | New use |  |
| --- | --- | --- | --- | --- |
|  | Females | Males | Females | Males |
|  | % change (95%CI) | % change (95%CI) | % change (95%CI) | % change (95%CI) |
| **Overall** | 2.2 (0.3, 4.2) | 0.3 (-2.1, 2.8) | 5.1 (-1.1, 12.2) | -1.5 (-9.2, 7.6) |
| **Age group, years** |  |  |  |  |
| 10 - 17 | 11.7 (4.1, 20.5) | 1.4 (-3.9, 7.2) | 15.6 (8.5, 23.7) | -5.0 (-13.6, 5.7) |
| 18 - 24 | 7.5 (2.3, 13.2) | <0.1 (-4.6, 5.1) | 12.1 (-2.6, 32.2) | -4.5 (-10.4, 2.3) |
| 25 - 49 | 2.7 (-0.7, 6.4) | 0.8 (-2.0, 3.6) | 3.9 (-4.1, 13.3) | -0.5 (-8.6, 9.2) |
| 50 - 64 | 1.4 (1.1, 1.8) | 0.1 (-1.8, 2.1) | 5.5 (-2.5, 15.0) | -1.4 (-12.5, 13.0) |
| 65+ | -0.5 (-1.5, 0.6) | -0.4 (-3.0, 2.3) | -0.3 (-4.7, 4.5) | -0.8 (-8.6, 8.4) |
| **Type of antidepressant** |  |  |  |  |
| SSRI | 2.3 (0.7, 4.0) | -0.8 (-2.6, 1.1) | 4.2 (-0.7, 9.6) | -4.7 (-10.8, 2.3) |
| Mirtazapine | 0.2 (-1.1, 1.5) | -0.2 (-2.2, 1.9) | 3.4 (-0.1, 7.0) | -1.2 (-7.6, 6.0) |
| TCA | 4.7 (-2.1, 11.8) | 2.3 (-7.0, 13.7) | 5.3 (-5.9, 19.4) | 1.4 (-14.6, 24.9) |
| SNRI | 1.2 (-1.4, 3.8) | 2.2 (0.1, 4.2) | 10.5 (-8.6, 39.7) | 8.4 (-12.4, 42.3) |
| Other/ MAOI | -6.4 (-7.9, -4.9) | -6.0 (-14.1, 3.9) | -11.3 (-27.3, 13.8) | -10.6 (-29.5, 22.0) |

\*From March 2020 to February 2021. % change refers to relative change.
SSRI: selective serotonin reuptake inhibitors, TCA: tricyclic antidepressants, MAOI: monoamine oxidase inhibitors, SNRI: serotonin-noradrenaline reuptake inhibitors

**Figure 1.**
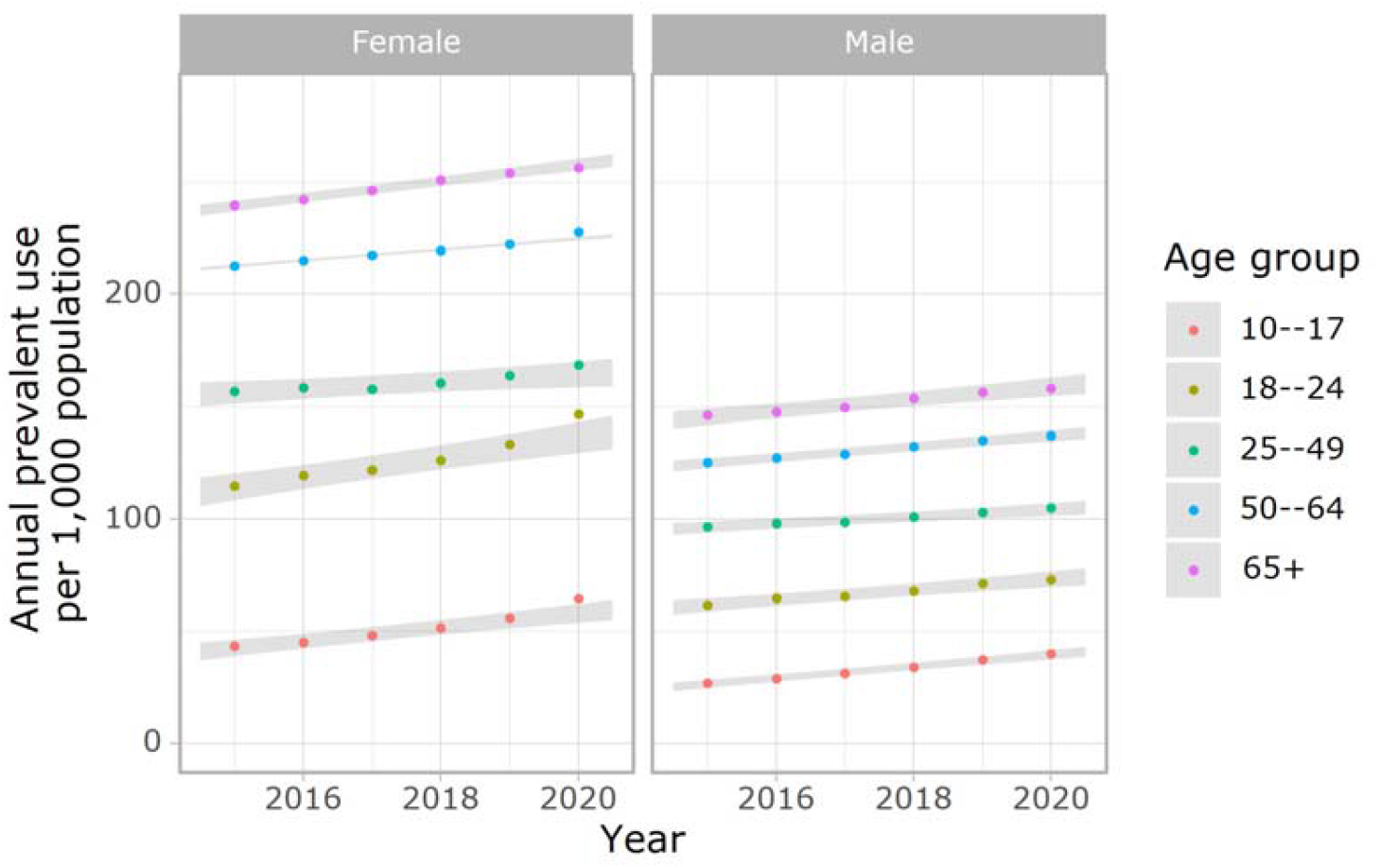
Trends in prevalent antidepressant use according to age and sex, 2015-2020. Note: Each year shown in the graph includes data from one year range (e.g., the 2020 data point represents data spanning from 2020-2021). Shaded areas represent 95% prediction intervals of trends. Point estimates within shaded area in 2020 represent non-statistically significant changes during the COVID-19 pandemic relative to predicted values.

The overall incidence of antidepressant use did not change discernibly during the first year of the COVID-19 pandemic (+2.3%, 95%CI −4.4%, 10.1%) (Table 2, Figure 2). But among adolescent females we observed a large and higher than expected increase in antidepressant initiation (+15.6% 95%CI 8.5%, 23.7%), corresponding to an excess of 5,068 (95%CI 2,956, 7,173) persons.

**Figure 2.**
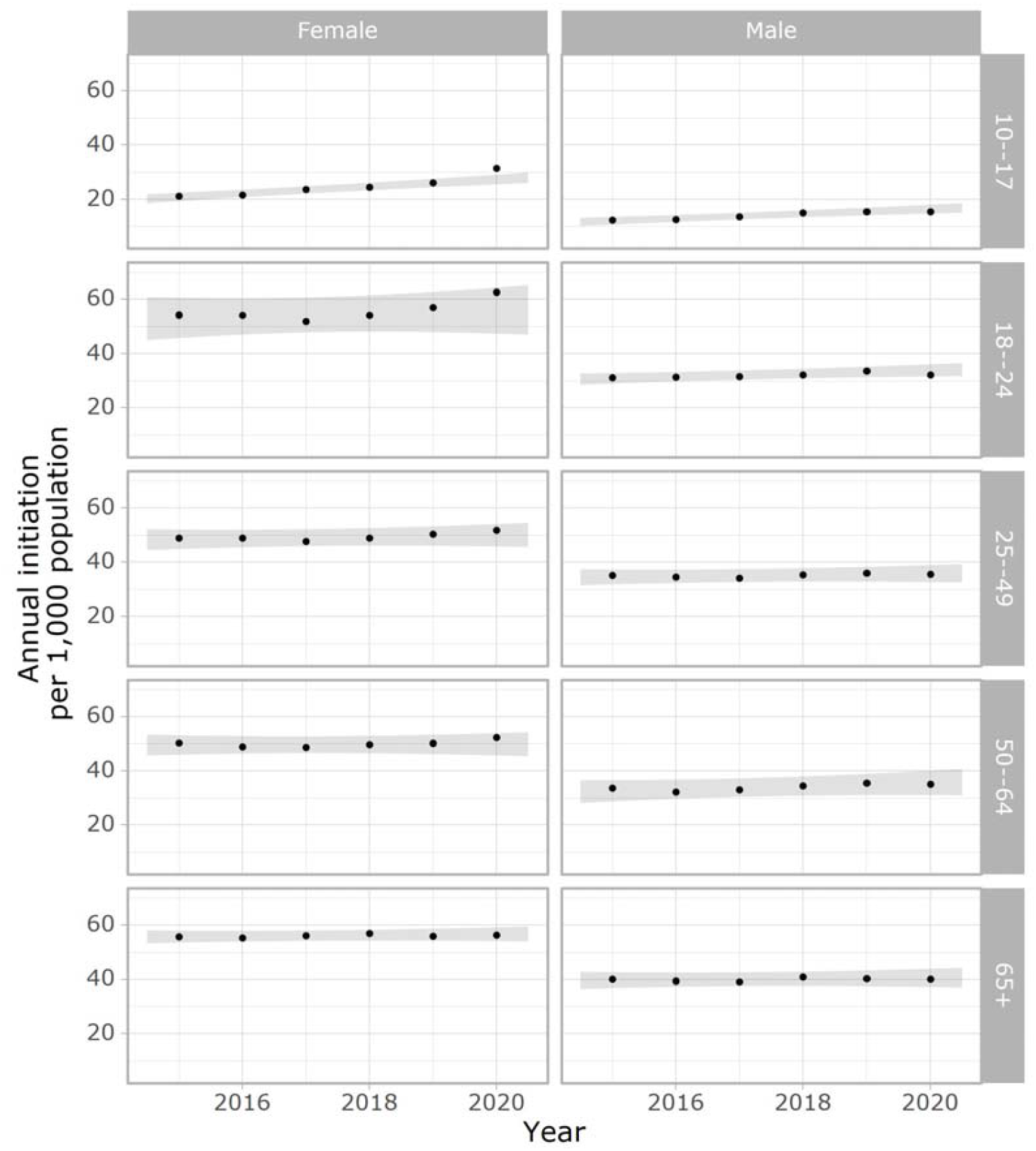
Trends in new antidepressant use according to age and sex, 2015-2020. Note: Each year shown in the graph includes data from one year range (e.g., the 2020 data point represents data spanning from 2020-2021). Shaded areas represent 95% prediction intervals of trends. Point estimates within shaded area in 2020 represent non-statistically significant changes during the COVID-19 pandemic relative to predicted values.

Sensitivity analysis: Analysis based on the alternative population estimates for 2020 yielded an even greater increase in prevalent antidepressant use among females (+11.2%, 95%CI 5.0%, 18.3%) and males (+4.1, 95% −1.2%, 10.0%) aged 18 to 24 years (See supplementary table 1). As expected, results did not change substantially for the other age groups, nor for the number of antidepressants dispensed per 1,000 population (+3.0%, 95% 0.7%, 5.3%) when compared to the base case scenario.

## Discussion

In this Australia-wide study, we found increasing trends in prevalent and new antidepressant use, suggesting a high and increasing burden of treated depression and anxiety. Consistent with a higher burden and diagnosis rates of these conditions among females, prevalent antidepressant use among women was approximately 1.7 times higher than men; females across all age groups had higher use of antidepressants compared to males ^7 8 23-25^. Our results also identified that females, especially adolescents, had the highest relative increases in antidepressant use during the first year of the COVID-19 pandemic.

Antidepressant use is increasing worldwide, both in terms of dispensed volume ^1^ and number of people using these medicines ^8 23-25^. Our prevalence estimates (9.3%-10.4% among men and 15.9%-17.6% among women) are similar to population-wide estimates among adults in the United States (13.8%) and New Zealand (12.6%) ^8 25^, although results are not directly comparable due to differences in ascertainment methods (e.g., self-report vs dispensing data) and periods evaluated. Our estimates, including the entire population, were substantially lower than previously reported in Australia for people receiving government benefits (21.6%-25.2% between 2007 and 2015) ^3^. This was expected, given people receiving government benefits are more likely to be older with higher rates of medicine use compared to the general population ^2^. We observed a lower rate of increase in the antidepressant treatment initiation than prevalent use, suggesting durations of antidepressant treatment have increased ^3 4^. Treatment guidelines recommend antidepressant use for at least 6-12 months, with limited evidence of benefits in use for periods longer than 3 years ^26^. Still, the average treatment duration in Australia is about four years, with lower durations among younger people and peaking at five years among those aged 55 to 64 years ^4^.

Of note are increases in antidepressant use among adolescents, especially new antidepressant use among females. The upward trend in antidepressant use among adolescents has been previously reported in Australia, with fluoxetine the most commonly dispensed for people aged 10 to 19 years ^9^. Antidepressant treatment has been associated with increased suicidal ideation, particularly in the first 2-4 weeks of treatment ^27^, with SSRIs used commonly for self-poisoning among adolescents ^11^. For those reasons, regulators in Australia and elsewhere warn about the risks of clinical worsening and suicide in approved prescribing information, specifically mentioning children, adolescents and young adults. Clinical guidelines also recommend prescribers to initiate the lowest possible dose of antidepressants, keeping the minimum supply needed for treatment when prescribing for children and adolescents ^28^.

Changes in mental health during COVID-19 may be reflected in patterns of antidepressant use in the population. Several jurisdictions worldwide reported stockpiling of antidepressants, in some cases followed by a sustained increase in use over several months ^18 29-34^. In our study, we observed females had higher than expected increases in antidepressant use in the first year of the COVID-19 pandemic relative to predicted rates, especially female adolescents, and female young adults, populations already on a trajectory of increased use of antidepressants prior to the pandemic. This finding is consistent with the increased depressive symptoms and psychological distress among young people during the first waves of COVID-19 in Australia ^18 35^, but in contrast to our findings, sex differences were not observed in younger age groups ^35^. The higher distress among adolescents and young adults during the COVID-19 pandemic is possibly due to more significant disruptions in daily routines and social interactions (e.g., interrupted education and workplace training and increased loneliness) and higher job insecurity in this population ^36 37^.

Anticipating the adverse impacts of COVID-19 restrictions on mental health, the Australian government launched several initiatives to maintain and expand access to mental health services, including temporary telehealth medical consultations, digital prescriptions, psychological therapy sessions, helplines services, and pop up clinics ^38 39^ Thus, increases in antidepressant use observed in our study may be a consequence of both increased need and availability of medical services. As observed in other countries, with different levels of COVID-19 infection rates and health care systems, the distress associated with the first year of the COVID-19 pandemic was not accompanied by higher suicide rates ^40^. In fact, the age-standardised death rate for suicide in Australia in 2020 (12.1/100,000 people) was 6.2% lower than in 2019 ^41^. This reduction likely reflects the combined effect of multiple factors such bolstering mental health services, changes in help-seeking patterns (e.g., increased use of psychological consultations, help lines and self-support tools), financial support packages implemented by the government, the initial community response of mutual support and an increased sense of belonging, among other factors ^42^. That aside, there is also evidence that suicidal and self-harm presentations by younger people (<18 years of age) increased while rates in other age groups did not ^42^. Taken as a whole, these findings highlight the need to monitor patterns of treatment, self-harm presentations, and suicide rates among subgroups of the population and target health initiatives accordingly.

### Strengths and limitations

Our study is a contemporary, national perspective on patterns of antidepressant use by age and sex, generating insights for mental health services planning. Our analysis of the first year of COVID-19 in Australia identifies the population subgroups most affected by the pandemic, looking beyond the initial stockpiling effects in the early months of the pandemic and considering the impact of population estimate uncertainty on rates of antidepressant use.

The main limitations of this study are related to the scope of PBS data. The lack of information on reasons for prescription means antidepressants could have been prescribed for conditions other than anxiety and depression. This is particularly relevant for SNRIs and TCAs, commonly used to treat pain. In addition, since PBS data do not include private and inpatient prescriptions in public hospitals, we underestimate the population-level use of antidepressants. Additionally, due to the drug utilisation design of the study, we provide limited insight into the course of treatment at an individual rather than population level. Finally, our findings are likely only generalisable to other jurisdictions where the circulation of SARS-CoV-2 was limited. We also expect differences in later Australian data due to the higher case numbers and extended lockdown in 2021.

## Conclusion

Antidepressant use continues to grow in Australia overall, with sustained high use among females and increased new use among adolescents despite safety concerns. The impact of the first year of the COVID-19 pandemic in antidepressant use was age and sex specific, likely reflecting an increased mental health burden among females, particularly adolescent females. Reasons for these discrepancies require further investigation.

## Data Availability

Aggregate Pharmaceutical Benefits Scheme (PBS) claims data are publicly available at https://www.pbs.gov.au/info/statistics/dos-and-dop/dos-and-dop. The PBS 10% sample data were used under license from the Australian Government Services Australia. Access to these data by other individuals or authorities is not permitted without the express permission of the approving human research ethics committees and data custodians.

## Acknowledgements

The authors thank Services Australia for providing the data.

## Appendix

**Supplementary box 1.** Antidepressant class classification.

| Class | Anatomical Therapeutic Chemical (ATC) Classification System codes | Generic name |
| --- | --- | --- |
| Selective serotonin reuptake inhibitors (SSRI) | N06AB02–N06AB10 | Citalopram, escitalopram, fluoxetine, fluvoxamine, paroxetine, sertraline |
| Mirtazapine | N06AX11 | Mirtazapine |
| Serotonin-Noradrenaline Reuptake Inhibitors (SNRI) | N06AX16, N06AX21, N06AX23 | Venlafaxine, duloxetine, desvenlafaxine |
| Tricyclic Antidepressants (TCA) | N06AA01–N06AA23 | Amitriptyline, clomipramine, desipramine, dothiepin, doxepin, imipramine, nortriptyline, trimipramine |
| Monoamine oxidase inhibitors (MAOI) | N06AF01–N06AF06<br>N06AG02–N06AG03 | Moclobemide, phenelzine, tranylcypromine |
| Other antidepressants | N06AX03, N06AX06, N06AX18 | Mianserin, nefazodone, reboxetine |

**Table S1.**
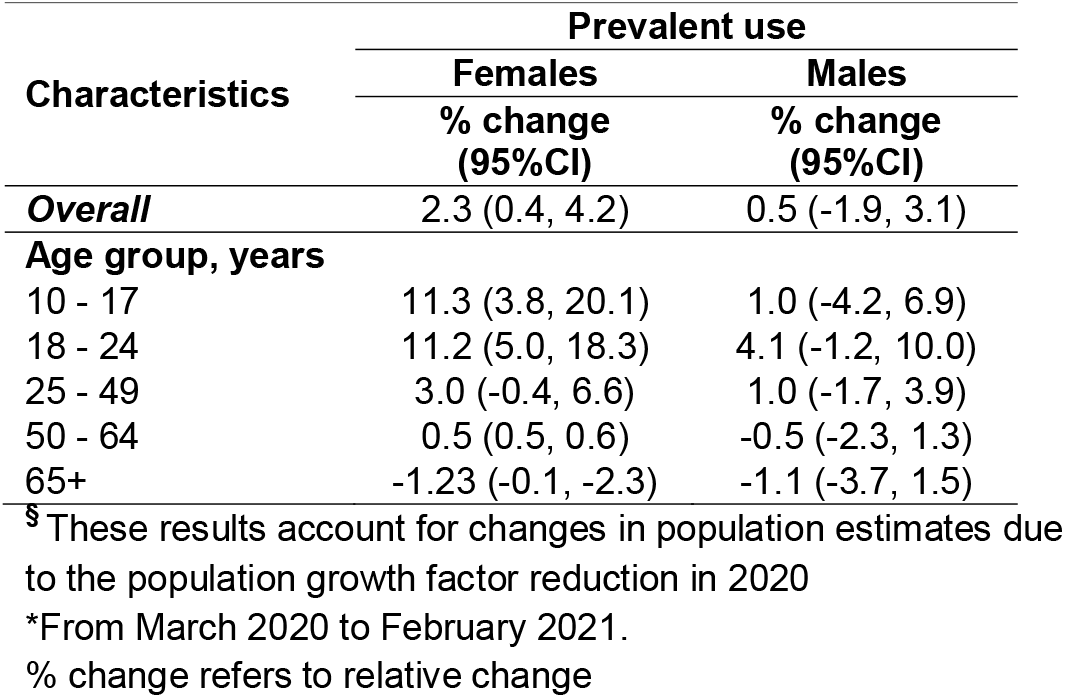
Results from the sensitivity analysis^§^ on changes in prevalent antidepressant use during the first year of COVID-19* according to sex and age.

| Characteristics | Prevalent use |  |
| --- | --- | --- |
|  | Females | Males |
|  | % change (95%CI) | % change (95%CI) |
| **Overall** | 2.3 (0.4, 4.2) | 0.5 (-1.9, 3.1) |
| **Age group, years** |  |  |
| 10 - 17 | 11.3 (3.8, 20.1) | 1.0 (-4.2, 6.9) |
| 18 - 24 | 11.2 (5.0, 18.3) | 4.1 (-1.2, 10.0) |
| 25 - 49 | 3.0 (-0.4, 6.6) | 1.0 (-1.7, 3.9) |
| 50 - 64 | 0.5 (0.5, 0.6) | -0.5 (-2.3, 1.3) |
| 65+ | -1.23 (-0.1, -2.3) | -1.1 (-3.7, 1.5) |

<sup>s</sup> These results account for changes in population estimates due to the population growth factor reduction in 2020
\*From March 2020 to February 2021.
% change refers to relative change

